# Antibiotic prescribing records in two UK primary care electronic health record systems. Comparison of the CPRD GOLD and CPRD Aurum databases

**DOI:** 10.1101/2020.03.07.20028290

**Authors:** Martin C Gulliford, Xiaohui Sun, Thamina Anjuman, Eleanor Yelland, Tarita Murray-Thomas

**Affiliations:** King’s College London, School of Population Health and Environmental Sciences, London, UK; National Institute for Health Research Biomedical Research Centre at Guy’s and St Thomas’ National Health Service Foundation Trust, London UK; Clinical Practice Research Datalink, Medicines and Healthcare Products Regulatory Agency, London, UK

**Keywords:** primary care, electronic healthcare records, antibiotics, infections

## Abstract

**Objective:** We evaluated whether recording of antibiotic prescribing across two primary care electronic health record (EHR) systems is similar. Data were analysed from the Clinical Practice Research Datalink (CPRD) databases: CPRD GOLD (Vision data) and CPRD Aurum (EMIS data).

**Methods:** Cohorts of patients were randomly sampled from both databases, stratifying by general practice, age-group and gender. All antibiotic prescriptions in 2017 were identified. Age- and sex-standardised antibiotic prescribing rates per 1,000 person years were calculated. Prescribing of individual antibiotic products and associated medical diagnosis recorded on the same date was also evaluated. English CPRD GOLD general practices were analysed as a subgroup, because all CPRD Aurum practices sampled were in England.

**Results:** There were 101,360 antibiotic prescriptions among 158,305 sampled patients at 883 CPRD Aurum practices, and 112,931 prescriptions among 160,394 sampled patients at 290 CPRD GOLD practices. The age- and sex-standardised antibiotic prescribing rate in 2017 was 512.6 (95% confidence interval 510.4 to 514.9) per 1,000 person years in CPRD Aurum and 584.3 (582.1 to 586.5) per 1,000 person years in CPRD GOLD [505.2 (501.6 to 508.9) per 1,000 person years if restricted to practices in England]. The 25 most frequently prescribed antibiotic products were similar in both databases. One or more medical codes were recorded on the same date as an antibiotic prescription for 72,989 (74%) prescriptions in CPRD Aurum, 84,756 (78%) in CPRD GOLD, and 28,471 (78%) for CPRD GOLD in England. Skin, respiratory and genito-urinary tract infections were recorded for 39,035 (40%) prescriptions in CPRD Aurum, 41,326 (38%) in CPRD GOLD, with 15,481 (42%) in English CPRD GOLD practices only.

**Conclusion:** Similar estimates for antibiotic prescribing and infection recording were found for both databases suggesting similar recording across EMIS and Vision systems. Future research on antimicrobial stewardship can be conducted in CPRD Aurum informed by previous results from CPRD GOLD. It may also be possible to combine CPRD GOLD and CPRD Aurum data in research on antibiotic prescribing.

## INTRODUCTION

Primary care electronic health records (EHRs) provide an important source of longitudinal population-based data for public health research and population health surveillance.(1) In the UK, several EHR databases collect de-identified data from general practices, which are responsible for providing a broad range of general medical services in the primary care setting. The CPRD GOLD database (1) and The Health Improvement Network (THIN) database (2) collect data from general practices that use the Vision practice system; data from EMIS practice systems have been historically collected by the QResearch database.(3) CPRD has now established a new database, CPRD Aurum (4) that also collects data from general practices using the EMIS practice system.

While the research community has nearly 30 years of experience of using Vision data from CPRD GOLD, with numerous studies reporting on data quality,(5, 6) less is known about the similarities and differences with data collected in the CPRD Aurum database. EHR systems may offer end-users a variety of options, and differing incentives, to code clinical data and record test results. It is therefore possible that analysis of Vision-derived data from CPRD GOLD and EMIS-derived data from CPRD Aurum may yield different findings when substantive research questions are addressed. However, the magnitude of any possible differences between these data sources are largely unexplored. This study aimed to compare results obtained from an analysis of CPRD GOLD and CPRD Aurum data for one exemplar, the recording of antibiotic prescribing.

Antibiotic prescribing in primary care has been the subject of increasing interest in recent years because of the growing awareness that unnecessary prescription of antibiotics in primary care is contributing to the problem of antimicrobial resistance.(7, 8) Research using CPRD GOLD (9, 10) and THIN, (11) showed that the antibiotic prescribing rate is between 500 and 600 antibiotic prescriptions per 1,000 patient years, with higher rates at the extremes of age. Nearly half of antibiotic prescriptions may be issued without a clear indication being recorded.(9, 11) The present analysis aimed to determine whether analysis of data from CPRD Aurum and CPRD GOLD provides similar estimates with respect to antibiotic prescription and recording.

## METHODS

### Data and participants

The CPRD GOLD database collects data from the four countries of the UK, with 30% of contributing practices located in England at the time of this study. The CPRD GOLD database has been well described (1) and the high quality of the data collected has been documented in many studies.(12) The October 2019 database release from which the study cohort was sampled for this analysis included data on 17.6 million patients, of whom 2.6 million were currently active The CPRD Aurum database is more recently established, and at the time of this study (June 2019 release) drew on data collected from general practices in England only, using the EMIS practice system.(4) The CPRD Aurum database from which patients were sampled included data on 883 general practices with 23.1 million patients, including 2.5 million currently active patients. The study required analysis of anonymised data, the protocol for the study received scientific and ethical approval from the CPRD Independent Scientific Advisory Committee (ISAC Protocol 19_110R).

A sample of 158,305 patients in CPRD Aurum was taken by randomly selecting ‘n’ patients from each stratum of general practice, gender and age-group. The value of n=9 was selected to provide an appropriate total sample size of just over 150,000. This sampling approach ensured that each general practice was equally represented in the analysis and that age-specific rates would be estimated with equal precision. Age was calculated as the difference between year of birth and 2017. Age-groups were categorised as 0-4, 5-14, 15-24, 25-34, 35-44, 45-54, 55-64, 65-74, 75-84 and 85 years and over.

A comparison cohort of patients was extracted from the October 2019 release of CPRD GOLD using the online interface. In this release, there were 290 general practices contributing data to CPRD GOLD throughout 2017. A sample of 160,394 patients was taken by randomly selecting n=30 patients from each stratum of general practice, gender and age-group. Patients were required to have at least 12 months of follow-up in the database estimated as the difference between the latest of their registration start date and 01/01/2017, and the earliest of registration end, practice last collection date, CPRD derived death date and 31/12/2017. General practices that migrated from Vision to EMIS practice systems during 2017 were excluded.

### Measures

We evaluated antibiotic prescribing for the year 2017 because this was the most recent complete year that we included in a larger study of antibiotic prescribing that we report elsewhere.(13) Antibiotics included all drugs from section 5.1 of the British National Formulary (BNF)(14) except anti-tuberculous, anti-lepromatous agents, and methenamine. The BNF includes the following categories of antibiotics: penicillins, cephalosporins (including carbapenems), tetracyclines, aminoglycosides, macrolides, clindamycin, sulfonamides (including combinations with trimethoprim), metronidazole and tinidazole, quinolones, drugs for urinary tract infection (nitrofurantoin), and other antibiotics. For CPRD GOLD, we employed a list of 2,627 antibiotic drugs that were identified from searches of the CPRD GOLD product dictionary browser made by all authors. Searches were made on the drug substance name, product name, BNF chapter and BNF codes. To identify the corresponding products in CPRD Aurum, dm+d codes (the prescribing codes from the National Health Service dictionary of medicines and devices) associated with individual product codes in the CPRD GOLD dictionary browser were mapped to the corresponding dm+d codes in the CPRD Aurum product dictionary browser. A more complete search of the CPRD Aurum product dictionary browser was additionally undertaken on term, product name and drug substance. We also conducted searches using approximate string matching (‘fuzzy matching’) to match the CPRD Aurum product name to the CPRD GOLD product name or drug substance name from the CPRD GOLD antibiotic code list. The ‘agrep’ command was used in the R program,(15) using the Levenshtein edit distance as a measure of approximateness. The resulting code list was edited manually, resulting in 896 CPRD Aurum product codes. CPRD Aurum product codes are up to 17 characters in length and the ‘bit64’ package in R was employed for data formatting and management.(16) Although more product codes were identified for the CPRD GOLD database, only 195 CPRD GOLD product codes for antibiotics, and 167 CPRD Aurum product codes were recorded during 2017.

Medical diagnoses were identified by searching the CPRD GOLD medical dictionary browser for Read terms and inspecting the associated Read chapter hierarchy. Separate codes lists were included for respiratory conditions, genito-urinary infections, skin infections and eye infections as reported previously.(9) The CPRD Aurum medical dictionary includes Read terms, Read codes and SNOMED codes. In order to utilise the same codes, lists developed for CPRD GOLD were subsequently mapped to CPRD Aurum by matching on Read codes. Evidence of infections were searched in the patient clinical and referral records in CPRD GOLD and in the observation tables in CPRD Aurum.

### Analysis

Age-specific rates were estimated with 95% confidence intervals from the Poisson distribution. Age- and sex-standardised rates, and associated 95% confidence intervals, were calculated per 1,000 person-years using the 2013 European Standard Population as reference. Differences between databases were evaluated using Bland-Altman plots.(17)

## RESULTS

### Comparison of antibiotic prescribing rates

In the CPRD Aurum sample, there were 158,305 participants from 883 general practices with 101,360 antibiotic prescriptions during 2017. In the CPRD GOLD sample there were 160,394 patients from 290 general practices with 112,931 antibiotic prescriptions during 2017. The age- and sex-standardised antibiotic prescribing rate was 512.6 (95% confidence interval 510.4 to 514.9) per 1,000 PYs in CPRD Aurum and 584.3 (582.1 to 586.5) per 1,000 PYs in CPRD GOLD. The rate for CPRD GOLD practices in England was 505.2 (501.6 to 508.9) per 1,000 PYs, which is similar to the rate observed in CPRD Aurum.

Figure 1 presents age- and sex-specific antibiotic prescribing rates for 2017. Antibiotic prescribing was higher in children under five years, decreased until the teenage years, increased again especially in women, before increasing steadily into older ages. This pattern of association was observed in both CPRD Aurum and CPRD GOLD but estimates for CPRD GOLD were generally slightly higher than for CPRD Aurum, but generally similar when restricted to CPRD GOLD general practices in England. The lower panel of Figure 1 provides a Bland-Altman plot that presents the difference (95% confidence interval) between CPRD GOLD and CPRD Aurum for all CPRD GOLD practices (blue) and CPRD GOLD practices in England (red). In men and women for all age groups, CPRD GOLD general practices generally had slightly higher antibiotic prescribing rates than CPRD Aurum, while CPRD GOLD general practices in England had similar antibiotic prescribing rates to CPRD Aurum.

**Figure 1:**
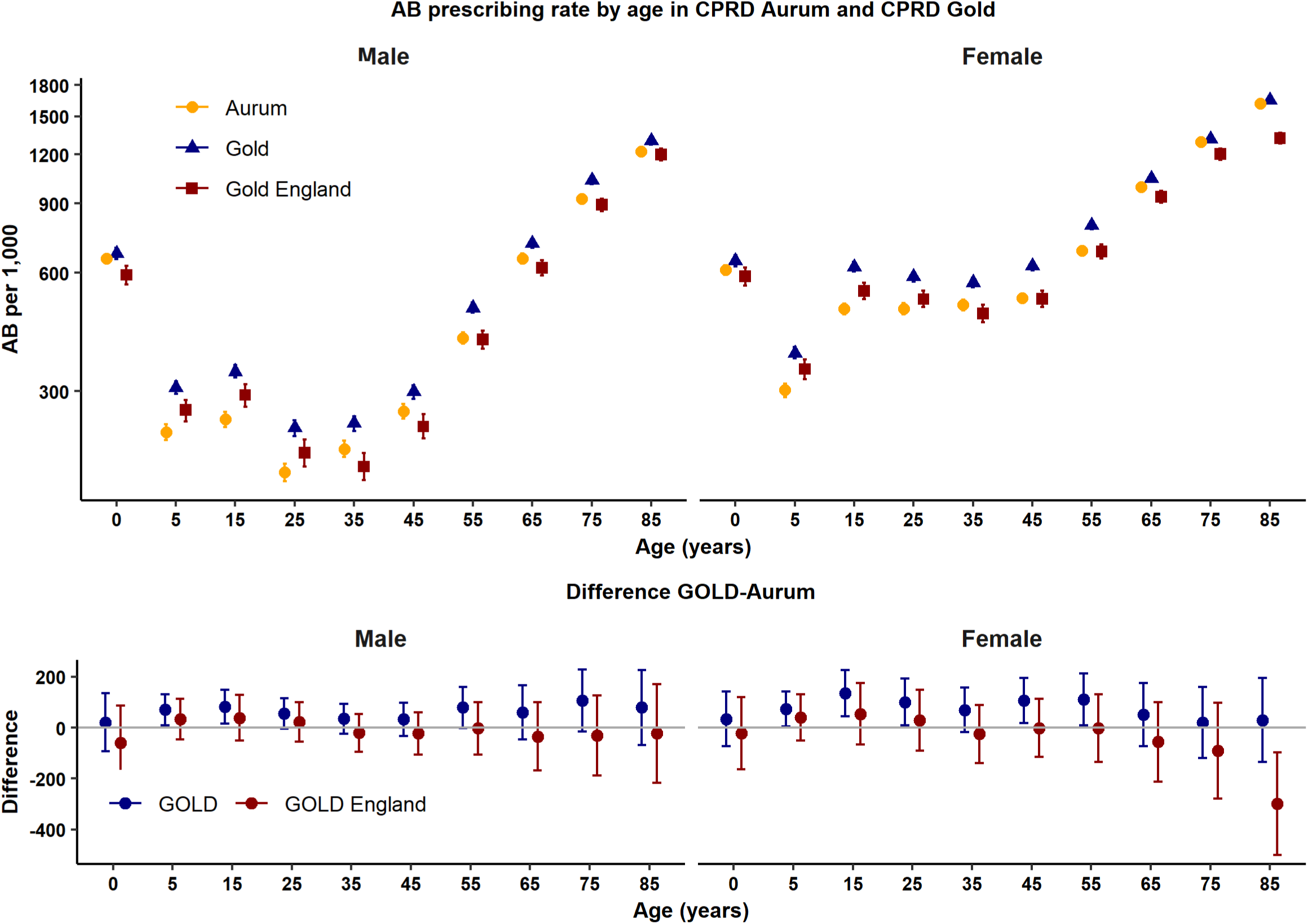
Antibiotic prescribing rates in CPRD Aurum and CPRD GOLD by age-group and sex.

### Most frequently prescribed products

Table 1 presents data for the 25 most frequently prescribed antibiotic products. In CPRD Aurum, amoxicillin 500mg capsules, doxycycline 100mg capsules, flucloxacillin 500mg capsules, trimethoprim 200mg tablets and nitrofurantoin 100mg modified-release capsules represented the five most frequently prescribed products, accounting for 45% of all antibiotic prescriptions. In CPRD GOLD, there were more prescriptions for trimethoprim and fewer prescriptions for nitrofurantoin, consequently clarithromycin 500 mg tablets and not nitrofurantoin appeared as the fifth ranked product. The same pattern was observed for CPRD GOLD practices in England, although trimethoprim comprised a smaller proportion of all prescriptions than in CPRD GOLD as a whole. Twenty-three of the 25 most frequently prescribed drugs in CPRD Aurum were also in the top 25 ranked prescriptions in CPRD GOLD general practices.

**Table 1:** Comparison of 25 most commonly prescribed antibiotic products in CPRD Aurum and CPRD GOLD.

| Product Name | CPRD Aurum |  |  | CPRD GOLD |  |  | CPRD GOLD England |  |  |
| --- | --- | --- | --- | --- | --- | --- | --- | --- | --- |
|  | Freq. | Percent | Rank | Freq. | Percent | Rank | Freq. | Percent | Rank |
| Amoxicillin 500mg capsules | 16,684 | 16.5 | 1 | 19,985 | 17.7 | 1 | 6,699 | 17.8 | 1 |
| Doxycycline 100mg capsules | 8,993 | 8.9 | 2 | 10,500 | 9.3 | 2 | 2,559 | 6.8 | 3 |
| Flucloxacillin 500mg capsules | 8,925 | 8.8 | 3 | 9,575 | 8.5 | 3 | 3,154 | 8.4 | 2 |
| Trimethoprim 200mg tablets | 5,931 | 5.9 | 4 | 9,064 | 8.0 | 4 | 2,277 | 6.1 | 4 |
| Nitrofurantoin 100mg modified-release capsules | 4,948 | 4.9 | 5 | 3,789 | 3.4 | 7 | 1,653 | 4.4 | 6 |
| Clarithromycin 500mg tablets | 4,656 | 4.6 | 6 | 4,889 | 4.3 | 5 | 1,842 | 4.9 | 5 |
| Phenoxymethylpenicillin 250mg tablets | 3,855 | 3.8 | 7 | 4,510 | 4.0 | 6 | 1,254 | 3.3 | 7 |
| Amoxicillin 250mg/5ml oral suspension sugar free | 3,030 | 3.0 | 8 | 3,645 | 3.2 | 8 | 1,065 | 2.8 | 9 |
| Co-amoxiclav 500mg/125mg tablets | 2,765 | 2.7 | 9 | 2,971 | 2.6 | 9 | 1,161 | 3.1 | 8 |
| Lymecycline 408mg capsules | 2,379 | 2.4 | 10 | 2,916 | 2.6 | 10 | 1,010 | 2.7 | 10 |
| Nitrofurantoin 50mg capsules | 2,295 | 2.3 | 11 | 2,821 | 2.5 | 11 | 848 | 2.3 | 12 |
| Trimethoprim 100mg tablets | 2,232 | 2.2 | 12 | 2,519 | 2.2 | 12 | 912 | 2.4 | 11 |
| Nitrofurantoin 50mg tablets | 2,169 | 2.1 | 13 | 1,539 | 1.4 | 18 | 610 | 1.6 | 17 |
| Amoxicillin 250mg/5ml oral suspension | 1,909 | 1.9 | 14 | 1,525 | 1.4 | 19 | 707 | 1.9 | 15 |
| Amoxicillin 125mg/5ml oral suspension sugar free | 1,802 | 1.8 | 15 | 990 | 0.9 | 25 | 369 | 1.0 | 25 |
| Azithromycin 250mg tablets | 1,602 | 1.6 | 16 | 1,306 | 1.2 | 22 | 373 | 1.0 | 24 |
| Erythromycin 250mg gastro-resistant tablets | 1,418 | 1.4 | 17 | 1,949 | 1.7 | 13 | 727 | 1.9 | 14 |
| Metronidazole 400mg tablets | 1,384 | 1.4 | 18 | 1,595 | 1.4 | 17 | 560 | 1.5 | 19 |
| Oxytetracycline 250mg tablets | 1,292 | 1.3 | 19 | 1,416 | 1.3 | 20 | 430 | 1.1 | 22 |
| Cefalexin 250mg capsules | 1,241 | 1.2 | 20 | 1,661 | 1.5 | 16 | 522 | 1.4 | 20 |
| Ciprofloxacin 500mg tablets | 1,236 | 1.2 | 21 | 1,724 | 1.5 | 15 | 662 | 1.8 | 16 |
| Amoxil 500mg capsules (GlaxoSmithKline UK Ltd) | 1,193 | 1.2 | 22 | 587 | 0.5 | 33 | 283 | 0.8 | 29 |
| Flucloxacillin 250mg capsules | 1,147 | 1.1 | 23 | 1,763 | 1.6 | 14 | 746 | 2.0 | 13 |
| Phenoxymethylpenicillin 125mg/5ml oral solution | 1,081 | 1.1 | 24 | 712 | 0.6 | 27 | 354 | 0.9 | 26 |
| Amoxicillin 250mg capsules | 1,073 | 1.1 | 25 | 1,397 | 1.2 | 21 | 575 | 1.5 | 18 |

### Recording of medical terms associated with prescriptions

Table 2 summarises data for recording of medical diagnostic codes on the same date as antibiotic prescriptions. Medical codes were recorded on the same date for 72,989 (74%) antibiotic prescriptions in CPRD Aurum, 84,756 (78%) antibiotic prescriptions in CPRD GOLD and 28,471 (78%) for CPRD GOLD in England. Infections of the skin, respiratory tract and genito-urinary tracts, accounted for 39,035 (40%) of CPRD Aurum prescriptions, 41,326 (38%) of CPRD GOLD prescriptions and 15,481 (42%) of CPRD GOLD prescriptions in England. All other medical codes accounted for 33,954 (34%) in CPRD Aurum, 43,430 (40%) in CPRD GOLD and 12,990 (36%) CPRD GOLD in England.

**Table 2:** Medical coding of antibiotic prescriptions. Figures are frequencies (percent of unique prescription dates).

|  | CPRD Aurum | CPRD GOLD | CPRD GOLD England |
| --- | --- | --- | --- |
| Number of prescription items | 101,360 | 112,931 | 37,551 |
| Number of prescriptions with unique date | 98,727 | 108,397 | 36,617 |
| Medical code recorded on same date | 72,989 (74.0) | 84,756 (78.2) | 28,471 (77.8) |
| No medical code recorded on same date | 25,738 (26.0) | 23,641 (21.8) | 8,146 (22.2) |
| Respiratory infection | 21,350 (21.6) | 26,005 (24.0) | 9,549 (26.1) |
| Genito-urinary infection | 11,126 (11.3) | 8,762 (8.1) | 3,315 (9.1) |
| Skin infection | 6,559 (6.6) | 6,559 (6.1) | 2,617 (7.1) |
| Other codes | 33,954 (34.4) | 43,430 (40.1) | 12,990 (35.5) |

## DISCUSSION

### Main findings

This analysis shows that antibiotic prescribing estimates from EMIS-derived data in CPRD Aurum are similar to those obtained through analysis of Vision-derived data in CPRD GOLD. This similarity includes the rates of antibiotic prescriptions for sub-groups of age and gender, the drug name and strength of antibiotic products prescribed, and the recording of medical diagnoses on the same day as the antibiotic prescription. We noted that antibiotic prescribing rates are generally higher in CPRD GOLD than in CPRD Aurum, but this was not the case when the CPRD GOLD sample was restricted to general practices in England. This implies that antibiotic prescribing may be higher in Wales, Scotland and Northern Ireland where there are no charges to patients for prescriptions. As well as slight differences in overall rates, we noted that drug choice might vary between databases. Trimethoprim prescribing was higher in CPRD GOLD than in CPRD Aurum. Nitrofurantoin has been recommended by Public Health England (18) as the drug of first choice for urinary tract infections in adults because of increasing antimicrobial resistance to trimethoprim. CPRD GOLD general practices in England were more similar to CPRD Aurum with respect to prescribing of trimethoprim and nitrofurantoin. It is likely that differences in clinical practice between England and the devolved administrations (Scotland, Wales and Northern Ireland) may be greater than the differences between EMIS and Vision practices within England.

Previous primary care database studies have revealed that antibiotic prescriptions are often issued without specific reasons being coded into electronic health records. A high proportion of antibiotic prescriptions in the THIN (11) and CPRD GOLD (9) databases are associated with either no code being recorded or non-specific codes being recorded. Electronic health records systems may offer users discretion over the recording of data items. We found that EMIS data included similar proportions of antibiotic prescriptions being associated with no codes, non-specific codes and codes for infection episodes.

### Strength and Limitations

One major strength of our study is that we used real world data from primary care to estimate rates of antibiotic prescribing. Using data from primary care is likely to provide a reliable picture of prescribing patterns given that about 80% of all antibiotic prescribing in the UK’s National Health Service take place in primary care.(18) We estimated the difference between CPRD Aurum and CPRD GOLD, as well as the difference between CPRD Aurum and CPRD GOLD in England. There were generally only small differences between CPRD Aurum and CPRD GOLD in England. We acknowledge that we could have obtained greater precision with larger samples, but the present approach was pragmatic and provided sufficiently precise estimates for age-specific rates. The community of CPRD researchers collectively have wide experience of compiling code lists for research in the CPRD GOLD database. Less experience is available for the CPRD Aurum database. We noted that CPRD Aurum product codes may be up to 17 characters in length and use of special programming features, such as the ‘bit64’ package in R,(16) are required in order to maintain data integrity. We completed extensive searches for product codes in order to identify antibiotic products. We identified a greater number of potential products from the CPRD GOLD data dictionary, but a generally similar number of antibiotic product codes were actually recorded in the two datasets during 2017. Searches in the CPRD Aurum product dictionary should be based on term, drug substance and product names as the BNF classification is less widely available in the CPRD Aurum product dictionary than in CPRD GOLD. It may also be possible to compare ‘dm+d’ codes from the dictionary of medicines and devices, which are now employed in both CPRD GOLD and CPRD Aurum product dictionaries. We mapped medical code sets between CPRD GOLD and CPRD Aurum by matching on Read codes to make a like-for-like comparison. The analysis shows that, for these conditions, use of the same Read codes gives similar results in CPRD Aurum and CPRD GOLD. There are some medical codes that are only employed in EMIS, which might be omitted through this process, and this merits further evaluation. Experience shows that in Read-coded data, the majority of events are associated with a small number of codes, consequently omission of infrequently used codes is seldom important; our main findings with respect to medical codes were consistent between databases. We also note that records of antibiotic prescribing do not indicate whether medicines were dispensed, whether they were taken, or whether they were taken by the patient they were prescribed to or by someone else. It is also possible that prescriptions recorded at out of hours visits, home visits or during attendance at residential care homes may be missing from the patient electronic record. Finally, the analysis undertaken was cross-sectional in nature and does not provide evidence about trends in antibiotic recording over time between CPRD GOLD and CPRD Aurum. We used data from July and October 2019 releases of CPRD Aurum and CPRD Gold respectively, it may be preferable to compare the same month’s releases but data for 2017 should be complete by 2019.

## Conclusion

This study finds that analysis of EMIS-derived data in CPRD Aurum gives generally similar estimates for antibiotic prescribing and infection recording to those reported for Vision-derived data in CPRD GOLD. CPRD GOLD includes general practices in Scotland, Wales and Northern Ireland which have slightly higher antibiotic prescribing than either EMIS or Vision general practices in England. Based on these results, we believe that future research studies can be conducted in CPRD Aurum, informed by previous results from CPRD GOLD or THIN. It may be possible to combine CPRD GOLD and CPRD Aurum data in research on antibiotic prescribing. However, further work is needed in other clinical topics and to understand the quality and completeness of information recorded in areas such as dosing regimen and treatment duration which is important in estimating treatment exposure in pharmacoepidemiology and pharmacovigilance research.

## Data Availability

Requests for access to data from the study should be addressed to. All proposals requesting data access will need to specify planned uses with approval of the study team and CPRD before data release.

## Data sources

The study is based on data from the Clinical Practice Research Datalink obtained under license from the UK Medicines and Healthcare products Regulatory Agency. However, the interpretation and conclusions contained in this report are those of the authors alone.

## Author Contributions

All authors contributed to the study design and definition of code lists. MG completed the analysis and drafted the paper. All authors reviewed and contributed to the final draft. MG is guarantor.

